# Molecular Heterogeneity in Early-Onset Colorectal Cancer: Pathway-Specific Insights in High-Risk Populations

**DOI:** 10.1101/2025.03.10.25323697

**Authors:** Cecilia Monge, Brigette Waldrup, Francisco G. Carranza, Enrique Velazquez-Villarreal

**Affiliations:** Center for Cancer Research, National Cancer Institute, Bethesda MD; City of Hope, Beckman Research Institute, Department of Integrative Translational Sciences, Duarte, CA; City of Hope Comprehensive Cancer Center, Duarte, CA

**Keywords:** early-onset colorectal cancer, cancer disparities, genetic mutations, precision medicine, WNT pathway, TGF-beta pathway, RTK/RAS pathway

## Abstract

**Background/Objectives:** The incidence of early-onset colorectal cancer (EOCRC), defined as diagnosis before age 50, has been rising at an alarming rate, with Hispanic/Latino (H/L) individuals experiencing the most significant increases in both incidence and mortality. Despite this growing public health concern, the molecular mechanisms driving EOCRC disparities remain poorly understood. Oncogenic pathways such as WNT, TGF-beta, and RTK/RAS are critical in colorectal cancer (CRC) progression, yet their specific roles in EOCRC across diverse populations have not been extensively studied. This research seeks to identify molecular alterations within these pathways by comparing EOCRC cases in H/L and non-Hispanic White (NHW) individuals. Furthermore, we explore the clinical significance of these findings to inform precision medicine strategies tailored to high-risk populations.

**Methods:** To investigate mutation frequencies in genes associated with the WNT, TGF-beta, and RTK/RAS pathways, we conducted a bioinformatics analysis using publicly available CRC datasets. The study cohort consisted of 3,412 patients, including 302 H/L and 3,110 NHW individuals. Patients were categorized based on age (EOCRC: <50 years; late-onset CRC [LOCRC]: ≥50 years) and population group (H/L vs. NHW) to assess variations in mutation prevalence. Statistical comparisons of mutation rates between groups were conducted using chi-squared tests, while Kaplan-Meier survival analysis was employed to evaluate overall survival differences associated with pathway alterations.

**Results:** Notable molecular distinctions in the RTK/RAS pathway were identified between EOCRC and LOCRC among H/L patients, with EOCRC exhibiting a lower frequency of RTK/RAS alterations compared to LOCRC (66.7% vs. 79.3%, p = 0.01). Within this pathway, mutations in CBL (p < 0.05) and NF1 (p < 0.05) were significantly more prevalent in EOCRC cases (5.8% vs. 1.2% and 11.6% vs. 3.7%, respectively), whereas BRAF mutations were notably less frequent in EOCRC than in LOCRC (5.1% vs. 18.3%, p < 0.05). Comparisons between EOCRC patients from H/L and NHW populations revealed distinct pathway-specific alterations that were more common in H/L individuals. These included RNF43 mutations (12.3% vs. 6.7%, p < 0.05) in the WNT pathway, BMPR1A mutations (5.1% vs. 1.8%, p < 0.05) in the TGF-beta pathway, and multiple RTK/RAS pathway alterations, such as MAPK3 (3.6% vs. 0.7%, p < 0.05), CBL (5.8% vs. 1.4%, p < 0.05), and NF1 (11.6% vs. 6.1%, p < 0.05). Survival analysis in H/L EOCRC patients did not reveal statistically significant differences based on pathway alterations. However, in NHW EOCRC patients, the presence of WNT pathway alterations was associated with significantly improved survival outcomes, suggesting potential ethnicity-specific prognostic implications.

**Conclusions:** This study highlights the substantial molecular heterogeneity present in EOCRC, particularly among high-risk populations. H/L EOCRC patients exhibited distinct genetic alterations, with a higher prevalence of CBL, NF1, RNF43, BMPR1A, and MAPK3 mutations compared to their NHW counterparts. Additionally, RTK/RAS pathway alterations were less frequent in EOCRC than in LOCRC. Despite these molecular differences, pathway alterations did not significantly impact survival outcomes in H/L EOCRC patients. However, in NHW EOCRC patients, the presence of WNT pathway alterations was associated with improved survival. These findings emphasize the necessity for further research to clarify the molecular mechanisms driving EOCRC disparities in high-risk populations and to inform precision medicine strategies for underrepresented groups.

## 1. Introduction

Colorectal cancer (CRC) remains a critical global health issue, ranking as the third most prevalent cancer and the second leading cause of cancer-related deaths worldwide (1). While advancements in screening and early detection have led to stable or declining CRC incidence in high-income countries, the incidence of early-onset colorectal cancer (EOCRC), diagnosed before the age of 50, has been increasing at an alarming rate (2–5). This rising trend is particularly concerning, as EOCRC is often associated with a more aggressive disease course and worse prognosis than late-onset colorectal cancer (LOCRC) (6). Since standard CRC screening guidelines typically recommend routine screening beginning at age 50, many EOCRC cases are detected at later stages, posing a significant public health challenge (7).

Among racial and ethnic groups, the H/L population in the United States has experienced the highest increase in EOCRC incidence and mortality (8, 9). Despite this growing burden, the molecular drivers of EOCRC in H/L individuals remain poorly characterized with limited sample sizes (10–12). The limited inclusion of H/L patients in large-scale genomic studies has hindered efforts to identify ethnicity-specific oncogenic mechanisms that may contribute to disparities in CRC incidence, progression, and clinical outcomes (13, 14). Addressing these disparities necessitates a deeper investigation into the molecular landscape of EOCRC in high-risk populations, particularly in relation to key oncogenic pathways.

Distinct molecular features have been observed in EOCRC, including higher microsatellite instability (MSI), increased tumor mutation burden, and elevated PD-L1 expression compared to LOCRC (3, 13, 14). Additionally, LINE-1 hypomethylation has been proposed as a unique biomarker of EOCRC (15). Comparative genomic analyses have highlighted significant differences in key oncogenic mutations between EOCRC and LOCRC, particularly in genes such as TP53, SMAD4, BRAF, NOTCH1, CTNNB1, APC, and KRAS (13, 16). However, few studies have specifically examined how these molecular alterations differ between H/L and non-Hispanic Whites (NHW) EOCRC patients, further underscoring the need for population-specific analyses.

Three critical signaling pathways implicated in CRC development and progression—the WNT, TGF-beta, and RTK/RAS pathways—play central roles in tumor initiation, invasion, and therapeutic resistance. The WNT pathway regulates β-catenin activation, which drives CRC progression and is commonly altered through mutations in APC, CTNNB1, and RNF43 (17, 18). Notably, WNT pathway activation is a hallmark of CRC, with mutations in APC observed in over 80% of cases (19). Studies suggest that EOCRC may exhibit unique WNT pathway alterations compared to LOCRC, with some analyses indicating a lower prevalence of WNT mutations in EOCRC (16), while others report elevated β-catenin activation (20, 21). A recent study demonstrated that EOCRC patients with WNT pathway alterations exhibited improved survival compared to those without, but this effect was observed primarily in NHW populations (10).

Similarly, the TGF-beta signaling pathway plays a crucial role in CRC progression, promoting epithelial-to-mesenchymal transition (EMT), immune evasion, and metastasis (22). TGF-beta pathway dysregulation is frequently observed in CRC, particularly through mutations in SMAD4 and BMPR1A, which have been associated with tumor aggressiveness and poor prognosis (23, 24). Genome-wide association studies (GWAS) have identified 16 genes within the TGF-beta pathway that are significantly associated with EOCRC (25). Among H/L patients, EOCRC-specific alterations in BMP7 and BMPR1A have been identified, suggesting population-specific differences in TGF-beta signaling (10).

The RTK/RAS pathway, which regulates cell proliferation, survival, and differentiation, is frequently altered in CRC. Mutations in KRAS, NRAS, and BRAF are among the most common genetic alterations in CRC, with KRAS mutations occurring in approximately 40% of cases (26). These mutations confer resistance to anti-EGFR therapies, a major challenge in CRC treatment (27). EOCRC patients exhibit higher frequencies of KRAS and NRAS mutations compared to LOCRC (7). However, recent studies suggest that RTK/RAS pathway alterations may be less prevalent in EOCRC compared to LOCRC in H/L patients, with an enrichment of mutations in CBL, NF1, and MAPK3 instead (10).

With the rising incidence of EOCRC among the H/L population and the limited understanding of ethnicity-specific oncogenic mechanisms, this study aims to explore the molecular heterogeneity of EOCRC in high-risk populations using a robust sample size. This approach seeks to address the historical underrepresentation of this group in clinical and genomic research, which is essential for advancing new technologies aimed at uncovering the molecular underpinnings of this cancer (28, 29). By examining pathway-specific alterations in WNT, TGF-beta, and RTK/RAS signaling, we aim to identify key molecular differences between EOCRC cases in H/L and NHW individuals (30, 31). Furthermore, we assess the clinical implications of these alterations to support the development of precision medicine strategies tailored to underrepresented populations. Gaining insight into the distinct molecular landscape of EOCRC in H/L patients is crucial for designing targeted therapeutic interventions and improving clinical outcomes in this high-risk group.

## 2. Materials and Methods

This study leveraged clinical and genomic data from 20 publicly available CRC datasets accessible through the cBioPortal database. The analyzed datasets encompassed colorectal adenocarcinoma, colon adenocarcinoma, and rectal adenocarcinoma, along with data from the GENIE BPC CRC v2.0-public dataset. To maintain a focus on primary tumor cases, datasets specifically examining metastatic CRC were excluded. Patient selection followed predefined inclusion criteria, requiring identification as Hispanic or Latino, Spanish, NOS; Hispanic, NOS; Latino, NOS; or individuals with a Mexican or Spanish surname. Additional filtering parameters ensured that only primary tumor cases were included, limiting selection to colorectal, colon, and rectal adenocarcinomas with confirmed adenocarcinoma, NOS histology, and allowing only one sample per patient.

After applying these criteria, four datasets—TCGA PanCancer Atlas, MSK Nat Commun 2022, MSK-CHORD, and GENIE BPC CRC—met the study’s requirements, resulting in a cohort of 302 H/L patients, including 138 EOCRC and 164 LOCRC cases. Similarly, 3,110 NHW patients (897 EOCRC and 2,213 LOCRC) were identified using identical inclusion criteria (Tables 1 & 2). Age at diagnosis was extracted from GENIE database clinical records. This study represents one of the most comprehensive investigations into WNT, TGF-beta, and RTK/RAS pathway alterations in an underserved population, offering crucial insights into the molecular disparities between EOCRC and LOCRC patients.

WNT, TGF-beta, and RTK/RAS pathway alterations were identified using established criteria, and patients were categorized into EOCRC, diagnosed before age 50, and LOCRC, diagnosed at age 50 or older. Further stratification was performed based on ethnicity (H/L vs. NHW) and the presence or absence of alterations in the aimed oncogenic pathways.

Table 3 details the distribution of these pathway alterations among EOCRC and LOCRC patients within the H/L cohort, illustrating their prevalence across different age groups.

Table 4 expands upon this analysis by comparing EOCRC cases between H/L and NHW patients, allowing for a comparative evaluation of pathway-specific molecular differences between ethnic groups. By incorporating these stratifications, this study provides a detailed molecular characterization of WNT, TGF-beta, and RTK/RAS pathway alterations, offering critical insights into potential ethnicity-related disparities that may inform precision medicine strategies for CRC.

Chi-square tests were employed to compare mutation frequencies across groups, assessing the independence of categorical variables and examining associations between age, ethnicity, and pathway alterations. To enhance the analysis, tumor samples were further categorized by anatomical location (colon vs. rectal adenocarcinoma), providing a more detailed evaluation of how tumor site interacts with ethnicity and pathway alterations. This stratification approach allowed for a more comprehensive investigation of patient heterogeneity, offering insights into potential variations in tumor biology and their implications for treatment responses.

Kaplan-Meier survival analysis was conducted to determine the prognostic impact of WNT, TGF-beta, and RTK/RAS pathway alterations on overall survival in different patient subgroups. Survival curves were generated to visualize variations in survival probability over time, comparing patients with and without these pathway alterations. The log-rank test was used to identify statistically significant differences between survival distributions, while median survival times and 95% confidence intervals (CIs) were calculated to ensure precise survival estimates. By leveraging large-scale genomic data, survival analyses, and subgroup comparisons, this study provides an in-depth evaluation of pathway-specific disruptions in EOCRC and LOCRC, particularly among H/L patients. These findings contribute to a broader understanding of ethnicity-specific molecular mechanisms underlying CRC.

## 3. Results

Using data from four cBioPortal projects that included ethnicity information, we established two cohorts: 302 H/L patients and 3,110 NHW patients. Within the H/L cohort, 27.5% of males and 18.2% of females were diagnosed with EOCRC before the age of 50, while 30.8% of males and 23.5% of females were diagnosed at age 50 or older (LOCRC). Comparatively, the NHW cohort had lower proportions of EOCRC cases, with 16.2% of males and 12.7% of females diagnosed before 50, whereas 38.9% of males and 32.3% of females were diagnosed with LOCRC (Table 1).

**Table 1.** Clinical and demographic profiles of Hispanic/Latino (H/L) and Non-Hispanic White (NHW) patient cohorts.

| Clinical Feature | H/L Cohort<br>n (%) | NHW Cohort<br>n (%) |
| --- | --- | --- |
| **Age Onset and Gender** |  |  |
| Early-Onset (< 50) Male | 83 (27.5%) | 503 (16.2%) |
| Early-Onset (< 50) Female | 55 (18.2%) | 394 (12.7%) |
| Late-Onset (≥ 50) Male | 93 (30.8%) | 1209 (38.9%) |
| Late-Onset (≥ 50) Female | 71 (23.5%) | 1004 (32.3%) |
| **Stage at Diagnosis** |  |  |
| Stage 1-3 | 98 (32.5%) | 965 (31.0%) |
| Stage 4 | 132 (43.7%) | 1131 (36.4%) |
| NA | 72 (23.8%) | 1014 (32.6%) |
| **Ethnicity** |  |  |
| Spanish NOS; Hispanic NOS, Latino NOS | 270 (89.4%) | 0 (0.0%) |
| Mexican (includes Chicano) | 28 (9.3%) | 0 (0.0%) |
| Other Spanish/Hispanic | 1 (0.3%) | 0 (0.0%) |
| Spanish surname only | 3 (1.0%) | 0 (0.0%) |
| Non-Spanish; Non-Hispanic | 0 (0.0%) | 3110 (100.0%) |

The gender distribution was relatively consistent across cohorts, with the H/L group comprising 55% males and 45% females, and the NHW group consisting of 56% males and 44% females. Regarding cancer stage at diagnosis, 32.5% of H/L patients were diagnosed at stages 0, I, II, or III, while 43.7% had stage IV disease. In the NHW cohort, 31.0% of patients were diagnosed at earlier stages (0–III), and 36.4% were diagnosed at stage IV. A notable proportion of cases had missing or unreported staging data, with 23.8% of H/L patients and 32.6% of NHW patients recorded as “NA” for stage at diagnosis.

Ethnicity classification within the H/L cohort showed that the majority (89.4%) identified as Spanish NOS, Hispanic NOS, or Latino NOS. Additionally, 9.3% of patients were classified as Mexican (including Chicano), while 1.3% identified as Other Spanish/Hispanic or were categorized based on a Spanish surname. In contrast, all NHW patients (100%) were classified as NHW, ensuring a clear distinction between the two groups for comparative analysis.

These demographic and clinical characteristics highlight notable differences in EOCRC incidence and disease stage at diagnosis between H/L and NHW populations, emphasizing potential disparities that could influence clinical outcomes.

A comparative analysis of clinical characteristics revealed significant differences between EOCRC and LOCRC in H/L patients, as well as between EOCRC H/L and EOCRC NHW patients (Table 2). The median age at diagnosis for EOCRC H/L patients was 41 years (IQR: 36–46), which was notably younger than the median age of 61 years (IQR: 55–69) for LOCRC H/L patients (p<0.05). Similarly, EOCRC H/L patients had a significantly lower median age at diagnosis compared to EOCRC NHW patients, whose median age was 42 years (IQR: 38–47) (p<0.05).

**Table 2.** Clinical characteristic differences between Hispanic/Latino (H/L) and Non-Hispanic White (NHW) cohorts based on ethnicity.

| Clinical Feature | Early-Onset HL<br>n (%) | Late-Onset HL<br>n (%) | p-value | Early-Onset HL<br>n (%) | Early-Onset NHW<br>n (%) | p-value |
| --- | --- | --- | --- | --- | --- | --- |
| Median Diagnosis Age (IQR) | 41 (36-46) | 61 (55-69) | < 0.05 | 41 (36-46) | 42 (38-47) | < 0.05 |
| Median Mutation Count* | 7 (5-10) | 8 (6-10) | > 0.05 | 7 (5-10) | 7 (5-10) | > 0.05 |
| RNF43 Mutation |  |  |  |  |  |  |
| Present | 17 (12.3%) | 22 (13.4%) | > 0.05 | 17 (12.3%) | 60 (6.7%) | <0.05 |
| Absent | 121 (87.7%) | 142 (86.6%) |  | 121 (87.7%) | 837 (93.3%) |  |
| BMPR1A Mutation |  |  |  |  |  |  |
| Present | 7 (5.1%) | 2 (1.2%) | > 0.05 | 7 (5.1%) | 16 (1.8%) | < 0.05 |
| Absent | 138 (100.0%) | 162 (98.8%) |  | 138 (100.0%) | 881 (98.2%) |  |
| BRAF Mutation |  |  |  |  |  |  |
| Present | 7 (5.1%) | 30 (18.3%) | < 0.05 | 7 (5.1%) | 67 (7.5%) | > 0.05 |
| Absent | 131 (94.9%) | 134 (81.7%) |  | 131 (94.9%) | 830 (92.5%) |  |
| MAPK3 Mutation |  |  |  |  |  |  |
| Present | 5 (3.6%) | 1 (0.6%) | > 0.05 | 5 (3.6%) | 6 (0.7%) | < 0.05 |
| Absent | 133 (96.4%) | 163 (99.4%) |  | 133 (96.4%) | 891 (99.3%) |  |
| CBL Mutation |  |  |  |  |  |  |
| Present | 8 (5.8%) | 2 (1.2%) | < 0.05 | 8 (5.8%) | 13 (1.4%) | < 0.05 |
| Absent | 130 (94.2%) | 162 (98.8%) |  | 130 (94.2%) | 884 (98.6%) |  |
| NF1 Mutation |  |  |  |  |  |  |
| Present | 16 (11.6%) | 6 (3.7%) | < 0.05 | 16 (11.6%) | 55 (6.1%) | < 0.05 |
| Absent | 122 (88.4%) | 158 (96.3%) |  | 122 (88.4%) | 842 (93.9%) |  |

\*LO HL: NA 1, EO NHW: NA 8

Mutation burden analysis showed that EOCRC H/L patients had a median mutation count of 7, slightly lower than the median of 8 observed in LOCRC H/L patients; however, this difference was not statistically significant (p>0.05). Likewise, EOCRC NHW patients exhibited a median mutation count of 7, which was similar to that of EOCRC H/L patients, with no significant difference between the two groups (p>0.05).

Pathway-specific analyses revealed significant differences in genetic alterations within the WNT, TGF-beta, and RTK/RAS pathways when comparing EOCRC and LOCRC in the H/L cohort. Notably, mutations in NF1 (11.6% vs. 3.7%, p<0.05), CBL (5.8% vs. 1.2%, p<0.05), and BRAF (5.1% vs. 18.3%, p<0.05) demonstrated significant variation between EOCRC and LOCRC cases. While BRAF mutations were significantly more prevalent in LOCRC cases, NF1 and CBL mutations were more frequently observed in EOCRC patients. Similarly, BMPR1A mutations were detected more often in EOCRC patients (5.1%) compared to LOCRC cases (1.2%), though this difference was not statistically significant (p>0.05).

A comparison between EOCRC H/L and NHW CRC patients further revealed ethnic-specific molecular differences. RNF43 (12.3% vs. 6.7%, p<0.05) from the WNT pathway, BMPR1A (5.1% vs. 1.8%, p<0.05) from the TGF-beta pathway, and MAPK3 (3.6% vs. 0.7%, p<0.05), CBL (5.8% vs. 1.4%, p<0.05), and NF1 (11.6% vs. 6.1%, p<0.05) from the RTK/RAS pathway were significantly more prevalent in EOCRC H/L patients compared to NHW patients. However, the frequency of BRAF mutations did not significantly differ between the two ethnic groups (5.1% vs. 7.5%, p>0.05). These findings highlight potential ethnicity-specific genomic differences that may play a role in CRC pathogenesis, warranting further investigation into their functional and clinical significance.

The variations in WNT, TGF-beta, and RTK/RAS pathway alterations observed between EOCRC and LOCRC, as well as between H/L and NHW EOCRC patients, emphasize the necessity for further research into their biological and clinical significance. A deeper understanding of these molecular differences could contribute to the development of targeted therapeutic approaches designed to reduce disparities in CRC outcomes across diverse ethnic groups.

Our analysis of genetic alterations in H/L individuals with EOCRC and LOCRC revealed no statistically significant differences in the prevalence of TGF-beta and WNT pathway alterations (Table 3). TGF-beta pathway mutations were identified in 34.1% of EOCRC cases and 33.5% of LOCRC cases, with identical absence rates of 65.9% and 66.5%, respectively (p = 1). Similarly, WNT pathway alterations were highly frequent in both groups, affecting 89.9% of EOCRC patients and 84.1% of LORCR patients, though the difference was not statistically significant (p = 0.1979). Notably, the absence of WNT pathway alterations was slightly higher in LOCRC patients (15.9%) compared to EOCRC patients (10.1%).

**Table 3.**
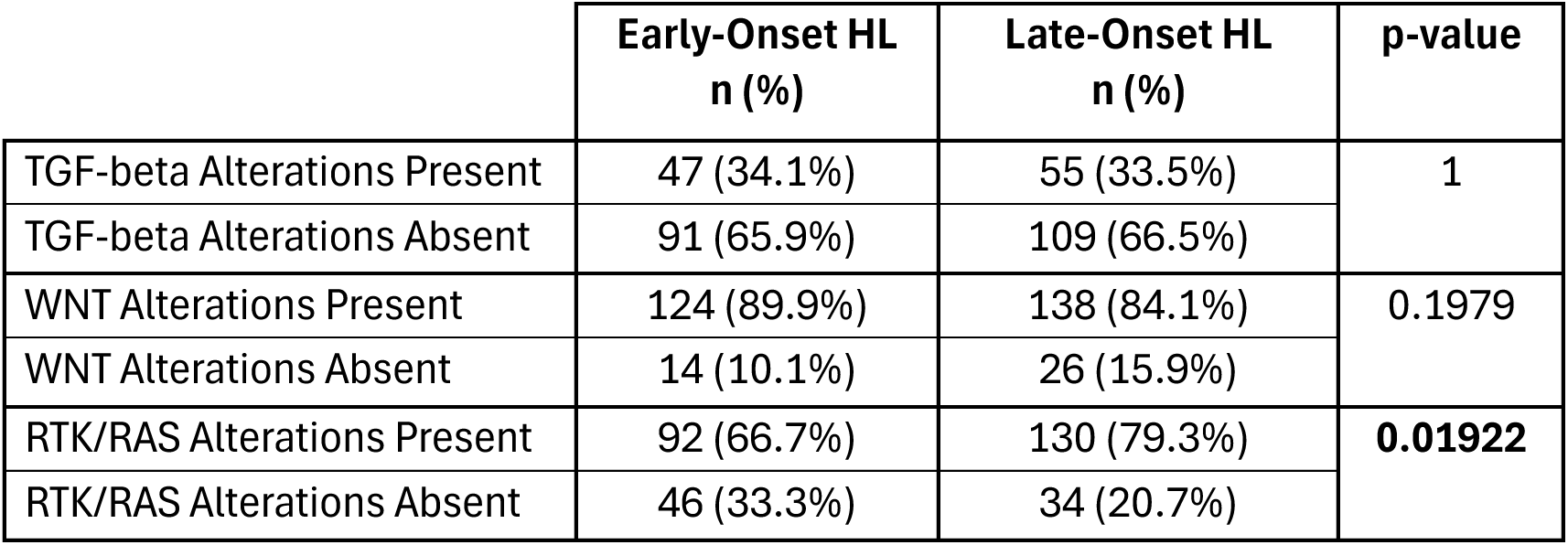
Frequency of WNT, TGF-beta, and RTK/RAS pathway alterations in Hispanic/Latino (H/L) patients diagnosed with either Early-Onset Colorectal Cancer (EOCRC) or Late-Onset Colorectal Cancer (LOCRC).

|  | Early-Onset HL<br>n (%) | Late-Onset HL<br>n (%) | p-value |
| --- | --- | --- | --- |
| TGF-beta Alterations Present | 47 (34.1%) | 55 (33.5%) | 1 |
| TGF-beta Alterations Absent | 91 (65.9%) | 109 (66.5%) |  |
| WNT Alterations Present | 124 (89.9%) | 138 (84.1%) | 0.1979 |
| WNT Alterations Absent | 14 (10.1%) | 26 (15.9%) |  |
| RTK/RAS Alterations Present | 92 (66.7%) | 130 (79.3%) | **0.01922** |
| RTK/RAS Alterations Absent | 46 (33.3%) | 34 (20.7%) |  |

In contrast, significant differences were observed in RTK/RAS pathway alterations between EOCRC and LOCRC patients. RTK/RAS mutations were detected in 66.7% of EOCRC cases, whereas they were more prevalent in LOCRC cases at 79.3% (p = 0.01922). Conversely, the proportion of patients without RTK/RAS alterations was higher in EOCRC (33.3%) compared to LOCRC (20.7%). These results indicate that while TGF-beta and WNT pathway mutations are consistently present in both age groups, RTK/RAS pathway alterations may have a greater influence on tumor development in LOCRC within the H/L population.

Additional research is needed to better understand the biological significance of RTK/RAS pathway variations between EOCRC and LOCRC patients. Moreover, examining how these pathways interact with other molecular drivers could offer valuable insights into CRC progression and identify potential therapeutic targets for this high-risk population.

Our analysis of genetic alterations in EOCRC among H/L and NHW individuals revealed significant differences in the frequency of TGF-beta pathway mutations, while no notable differences were observed in WNT and RTK/RAS pathway alterations (Table 4). TGF-beta pathway mutations were significantly more prevalent in EOCRC H/L patients (34.1%) compared to EOCRC NHW patients (25.5%) (p = 0.04489). In contrast, NHW individuals had a higher proportion of cases without TGF-beta pathway alterations (74.5%) compared to H/L individuals (65.9%).

**Table 4.**
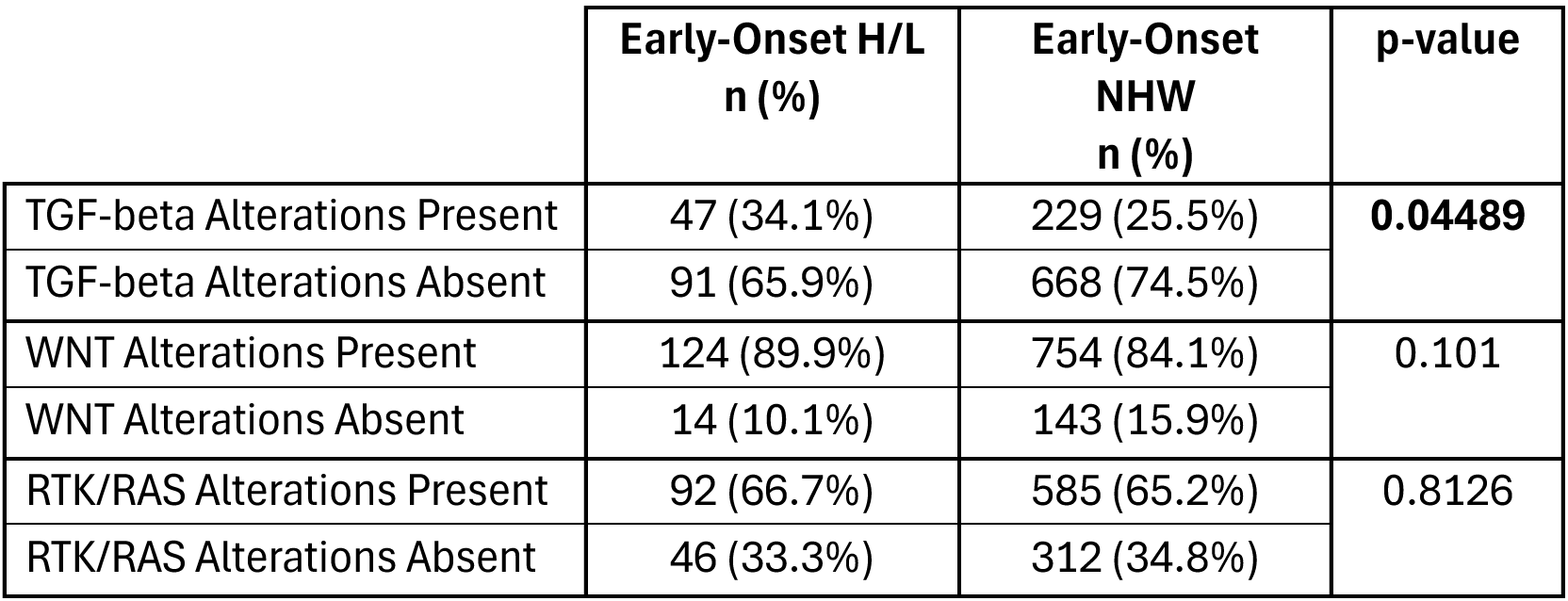
Frequency of WNT, TGF-beta, and RTK/RAS pathway alterations in Early-Onset Colorectal Cancer (EOCRC) among Hispanic/Latino (H/L) and Non-Hispanic White (NHW) patients.

Alterations in the WNT pathway were commonly observed in both EOCRC groups, affecting 89.9% of H/L patients and 84.1% of NHW patients. However, this difference did not reach statistical significance (p = 0.101). Conversely, the proportion of patients without WNT pathway alterations was slightly higher among NHW individuals (15.9%) compared to H/L individuals (10.1%).

RTK/RAS pathway alterations were detected in 66.7% of EOCRC H/L patients and 65.2% of EOCRC NHW patients, showing no significant difference between the two groups (p = 0.8126). Likewise, the proportion of patients without RTK/RAS pathway alterations remained similar, with 34.8% in the NHW group and 33.3% in the H/L group.

The results indicate that TGF-beta pathway alterations occur at a significantly higher frequency in EOCRC among H/L individuals, whereas WNT and RTK/RAS pathway mutations show no substantial differences between the two populations. To gain a deeper understanding of the clinical significance of these alterations and their potential role in precision medicine and targeted therapies for CRC, further research utilizing larger cohorts and functional studies is necessary.

Survival outcomes for EOCRC in H/L patients, analyzed using Kaplan-Meier and stratified by the presence or absence of TGF-beta pathway alterations (Figure 1), showed no statistically significant difference in overall survival (p = 0.74). The survival trajectories of both groups were largely overlapping, indicating that TGF-beta pathway mutations may have a minimal impact on survival outcomes within this cohort. Although patients with TGF-beta alterations showed a slightly lower survival probability during the initial months of follow-up, their survival curve eventually aligned with that of patients without alterations. The broad confidence intervals, especially at later time points, suggest variability in survival outcomes and the potential influence of additional molecular or clinical factors not accounted for in this analysis. These results imply that TGF-beta pathway alterations alone may not be strong prognostic markers in EOCRC H/L patients. Further research incorporating larger cohorts and multi-omic analyses is needed to explore whether interactions with other molecular pathways contribute to survival disparities in this high-risk population.

**Figure 1.**
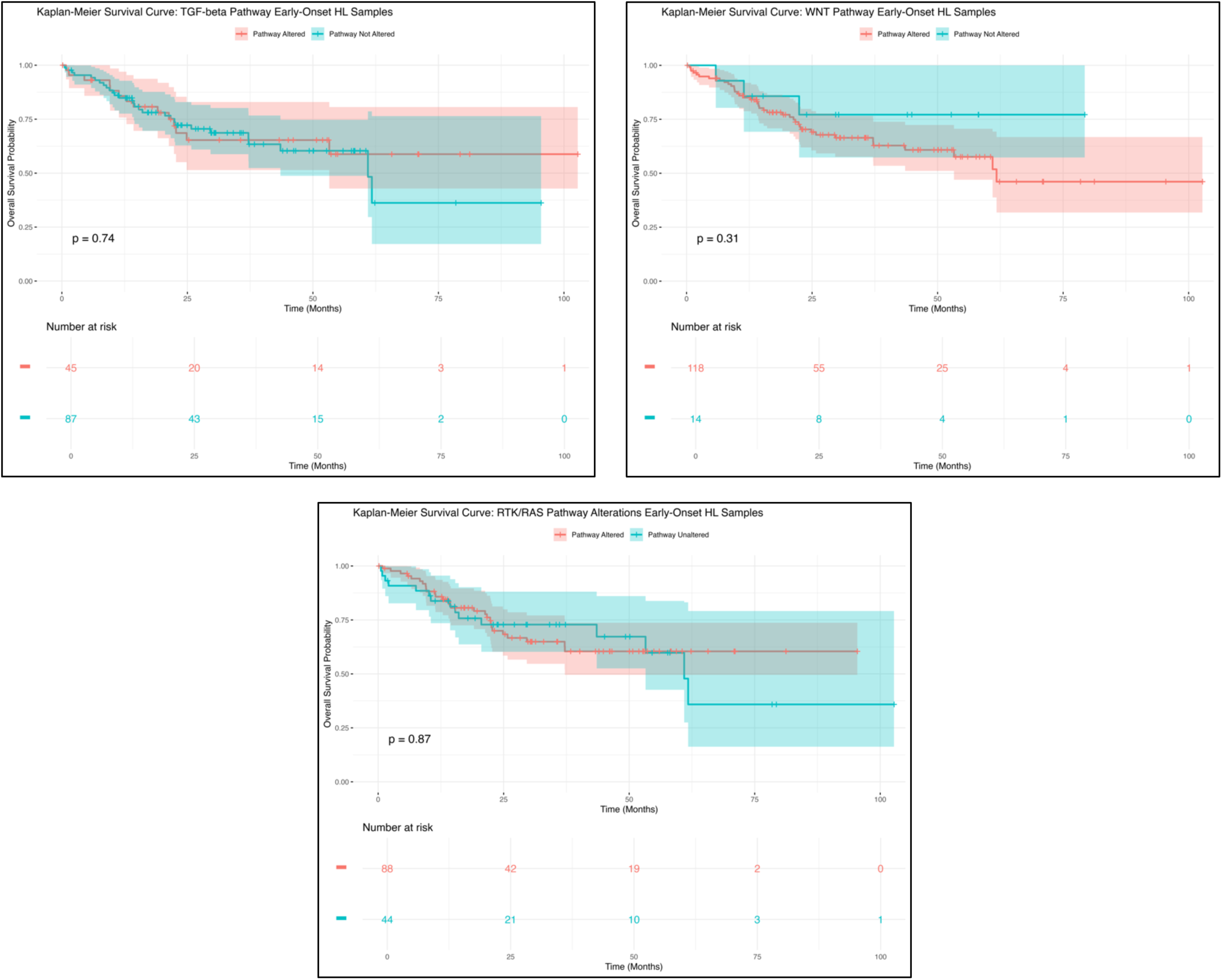
Kaplan-Meier overall survival curves for early-onset Hispanic/Latino patients, categorized based on the presence or absence of alterations in the TGF-beta (top left), WNT (top right), and RTK/RAS (bottom) pathways.

The analysis of overall survival in EOCRC H/L patients, based on the presence or absence of WNT pathway alterations (Figure 1), showed no statistically significant difference (p = 0.31). Although patients with WNT pathway alterations had slightly lower survival probabilities in the early months of follow-up, the survival curves later converged, indicating no clear link between WNT pathway mutations and long-term survival. Notably, these results differ from findings in NHW EOCRC patients, where WNT alterations were associated with better survival outcomes. The absence of statistical significance in the H/L cohort suggests that the prognostic role of WNT pathway mutations may vary across ethnic groups. Additional studies with larger sample sizes and functional investigations are needed to determine whether interactions with other oncogenic pathways contribute to survival differences in EOCRC among H/L patients.

Unlike the findings for TGF-beta and WNT pathway alterations, the survival analysis for EOCRC H/L patients based on RTK/RAS pathway alterations (Figure 1) indicated a possible trend toward worse survival outcomes in those with RTK/RAS mutations (p = 0.87). Patients with RTK/RAS alterations experienced an early decline in survival probability, maintaining lower survival rates throughout the follow-up period compared to those without such alterations. Conversely, patients without RTK/RAS pathway mutations exhibited a more stable survival probability over time. Although the difference did not reach statistical significance, the observed trend suggests that RTK/RAS mutations could play a role in survival disparities among EOCRC patients in the H/L population. These findings emphasize the need for further research to explore the impact of RTK/RAS pathway disruptions on CRC progression, particularly in underrepresented groups. Expanding sample sizes, conducting functional studies, and integrating multi-omic analyses will be essential to better understand the clinical relevance of RTK/RAS mutations and their implications for precision medicine in EOCRC among H/L patients.

Survival analysis using the Kaplan-Meier method for EOCRC NHW patients, categorized by the presence or absence of TGF-beta pathway alterations, showed no statistically significant difference in overall survival (p = 0.53) (Figure S1). The survival curves of both groups remained nearly identical, with minimal divergence, indicating that TGF-beta pathway alterations may not be a strong prognostic factor in NHW EOCRC. Although patients with TGF-beta alterations exhibited slightly lower survival probabilities in the early follow-up period, the curves eventually converged, suggesting no consistent survival trend. The wide confidence intervals and variability in survival estimates emphasize the need for further research with larger cohorts. Future investigations should examine whether TGF-beta alterations interact with other molecular or clinical factors that may impact disease progression and treatment response in NHW EOCRC patients.

Conversely, survival analysis using the Kaplan-Meier method for NHW EOCRC patients, categorized by the presence or absence of WNT pathway alterations, revealed a statistically significant association with improved overall survival (p = 0.0015) (Figure S1). Patients with WNT pathway mutations (red curve) consistently maintained higher survival probabilities across the follow-up period compared to those without mutations (blue curve), indicating a potential protective role of WNT alterations in this population. The clear separation between survival curves further supports a survival advantage for patients harboring WNT pathway mutations. While these findings underscore the prognostic significance of WNT alterations in NHW EOCRC, further research is necessary to examine potential confounding variables, elucidate the underlying biological mechanisms, and assess their impact on treatment response and long-term disease outcomes.

Kaplan-Meier survival analysis of NHW EOCRC patients, stratified by RTK/RAS pathway alterations, showed no statistically significant difference in overall survival (p = 0.18) (Figure S1). While the RTK/RAS-altered group (red curve) displayed a slight trend toward reduced survival compared to those without alterations (blue curve), the overlapping curves and wide confidence intervals indicate substantial variability in survival estimates. This variability may stem from factors such as sample size differences, molecular heterogeneity within the RTK/RAS-altered subgroup, and possible interactions with other oncogenic pathways. Although these findings do not strongly support RTK/RAS pathway alterations as a significant determinant of survival outcomes in NHW EOCRC patients, additional research with larger cohorts, functional molecular studies, and integrative pathway-based analyses is needed. Future investigations should assess how RTK/RAS alterations interact with other oncogenic pathways to clarify their clinical relevance in this population.

To evaluate potential age-related variations, the frequencies of gene alterations associated with the WNT, TGF-beta, and RTK/RAS pathways were examined in H/L patients diagnosed with EOCRC and LOCRC (Table S1). The findings revealed distinct differences in the prevalence of specific pathway alterations between the two groups.

In the WNT pathway, APC mutations were more prevalent among EOCRC patients (80.4%) than LOCRC patients (70.7%), though the difference was not statistically significant (p = 0.07022). Other key WNT pathway genes, such as AXIN1, AXIN2, and RNF43, displayed similar mutation frequencies between the two groups, with no significant variation. Likewise, no substantial differences in mutation rates were identified within the TGF-beta pathway between EOCRC and LOCRC patients. The most frequently altered gene in this pathway, SMAD4, had nearly identical mutation rates in both groups (14.5% in EOCRC vs. 14.6% in LOCRC, p = 1.0). Although BMPR1A mutations were slightly more common in EOCRC patients (5.1%) compared to LOCRC patients (1.2%), this difference did not reach statistical significance (p = 0.08853).

Within the RTK/RAS pathway, notable differences in gene alterations were observed between EOCRC and LOCRC patients. NF1 mutations were significantly more frequent in EOCRC cases (11.6%) compared to LOCRC cases (3.7%) (p = 0.01547). Likewise, CBL mutations were more prevalent in EOCRC patients (5.8%) than in LOCRC patients (1.2%) (p = 0.04765). Additionally, MAP2K1 mutations were found exclusively in EOCRC patients (3.6%) and were completely absent in LOCRC cases (p = 0.01914).

On the other hand, BRAF mutations were significantly more common in LOCRC patients (18.3%) compared to EOCRC patients (5.1%, p = 0.0009188), suggesting an age-related pattern in RTK/RAS pathway alterations.

The results indicate that certain RTK/RAS pathway alterations, including mutations in NF1, CBL, and MAP2K1, may have a greater impact on EOCRC, while BRAF mutations appear to be more prevalent in LOCRC. In contrast, the lack of significant variation in WNT and TGF-beta pathway alterations between EOCRC and LOCRC onset cases suggests that these pathways may drive CRC progression independently of age at diagnosis. Additional research is required to investigate the functional consequences of these molecular differences and their potential applications in precision oncology approaches for H/L CRC patients.

A comparative analysis of EOCRC between H/L and NHW patients revealed notable disparities in the mutation rates of key pathway genes (Table S2). Within the WNT pathway, RNF43 mutations were significantly more prevalent among EOCRC H/L patients (12.3%) compared to their NHW counterparts (6.7%, p = 0.02985). This suggests the possibility of ethnicity-specific variations in tumor suppressor gene alterations.

Within the TGF-beta pathway, BMPR1A mutations were notably more prevalent in EOCRC H/L patients (5.1%) than in their NHW counterparts (1.8%, p = 0.04443). In contrast, other key genes in this pathway, including TGFBR2 and SMAD4, did not exhibit statistically significant differences between the two groups. This finding implies that BMPR1A mutations may have a more prominent role in the tumorigenesis of EOCRC among H/L individuals.

In the RTK/RAS pathway, several genes displayed significantly higher mutation frequencies in EOCRC H/L patients compared to their NHW counterparts. Specifically, MAPK3 mutations were more prevalent in EOCRC H/L patients (3.6%) than in EOCRC NHW patients (0.7%, p = 0.006833). Similarly, NF1 mutations occurred at a significantly higher rate in H/L patients (11.6%) versus NHW patients (6.1%, p = 0.02907).

Additionally, CBL mutations were nearly four times more common in EOCRC H/L patients (5.8%) compared to NHW patients (1.4%, p = 0.002302). These findings suggest potential ethnic variations in RTK/RAS pathway alterations, which may play a role in CRC development and influence therapeutic responses.

On the other hand, mutation frequencies for KRAS, NRAS, and BRAF did not differ significantly between EOCRC H/L and NHW patients. This indicates that although certain RTK/RAS pathway genes exhibit ethnic-specific enrichment, the primary driver mutations appear to be consistent across both populations.

While WNT and TGF-beta pathway alterations show some ethnicity-specific variations, the most striking differences are observed in RTK/RAS pathway genes, especially NF1, CBL, and MAPK3. The significantly higher frequency of these mutations in EOCRC H/L patients highlights the necessity for further research into their functional impact on tumor development, resistance to therapy, and personalized treatment strategies for H/L CRC patients. Future investigations should focus on understanding how these molecular variations affect disease progression and response to targeted therapies across diverse racial and ethnic populations.

## 4. Discussion

CRC remains a significant public health challenge, with EOCRC exhibiting an alarming rise in incidence, particularly among H/L individuals. Despite this growing burden, the molecular mechanisms underlying EOCRC disparities remain poorly understood. This study provides a comprehensive analysis of WNT, TGF-beta, and RTK/RAS pathway alterations in EOCRC, revealing key differences between H/L and NHW patients. These findings contribute to the growing evidence that molecular heterogeneity plays a critical role in CRC pathogenesis and may inform precision medicine strategies tailored to underrepresented populations.

Our results highlight significant ethnic-specific differences in pathway alterations. Notably, RNF43, BMPR1A, MAPK3, NF1, and CBL mutations were significantly more prevalent in EOCRC H/L patients compared to their NHW counterparts. RNF43 mutations, a critical regulator of WNT signaling, were observed in 12.3% of EOCRC H/L patients compared to 6.7% in NHW patients (p < 0.05), suggesting potential differences in WNT pathway dysregulation across ethnic groups. Similarly, BMPR1A mutations, implicated in TGF-beta signaling, were significantly enriched in EOCRC H/L patients (5.1% vs. 1.8%, p < 0.05), reinforcing prior findings that alterations in this pathway may contribute to CRC progression in H/L individuals (5).

Within the RTK/RAS pathway, MAPK3 (3.6% vs. 0.7%, p < 0.01), NF1 (11.6% vs. 6.1%, p < 0.05), and CBL (5.8% vs. 1.4%, p < 0.01) mutations were significantly more frequent in EOCRC H/L patients than in NHW patients. The enrichment of NF1 mutations in EOCRC H/L patients is particularly intriguing, as NF1 loss has been associated with resistance to targeted therapies in CRC and other malignancies (6, 7). These findings suggest that ethnic-specific RTK/RAS pathway alterations may influence tumor behavior and treatment response in EOCRC, emphasizing the need for further studies evaluating the functional impact of these mutations.

Comparisons between EOCRC and LOCRC within the H/L cohort revealed key distinctions in molecular alterations. While WNT and TGF-beta pathway mutations occurred at comparable frequencies across age groups, RTK/RAS pathway alterations exhibited significant variation. NF1 (11.6% vs. 3.7%, p < 0.05), CBL (5.8% vs. 1.2%, p < 0.05), and MAP2K1 (3.6% vs. 0.0%, p < 0.05) mutations were significantly more prevalent in EOCRC H/L patients, suggesting a potential role for RTK/RAS dysregulation in EOCRC. Conversely, BRAF mutations were markedly higher in LOCRC patients (18.3% vs. 5.1%, p < 0.001), consistent with previous reports linking BRAF alterations to LOCRC and poor prognosis (8).

These findings support the notion that EOCRC and LOCRC may represent distinct molecular subtypes with unique oncogenic drivers. While NF1 and CBL mutations may contribute to tumor initiation in EOCRC, BRAF-driven signaling appears to play a more prominent role in LOCRC. The functional implications of these differences warrant further investigation, particularly in the context of targeted therapy development for EOCRC patients.

Kaplan-Meier survival analysis revealed significant differences in the impact of pathway alterations across ethnic groups. In EOCRC NHW patients, WNT pathway alterations were associated with improved survival (p = 0.0015), suggesting a potential protective role for WNT signaling mutations in this population. This aligns with prior studies indicating that APC mutations, a hallmark of WNT pathway activation, may be associated with better clinical outcomes in CRC (9). However, this survival advantage was not observed in EOCRC H/L patients, where WNT pathway mutations had no significant impact on overall survival (p = 0.31). These findings highlight potential ethnic-specific differences in the prognostic relevance of WNT signaling alterations, emphasizing the need for further investigation.

TGF-beta pathway alterations, in contrast, did not significantly impact survival outcomes in either EOCRC H/L (p = 0.74) or NHW (p = 0.53) patients. The absence of a strong prognostic association suggests that TGF-beta mutations alone may not drive survival differences in EOCRC, though their interactions with other molecular pathways remain an important area of study.

RTK/RAS pathway alterations demonstrated a trend toward poorer survival outcomes in EOCRC H/L patients (p = 0.087), though this did not reach statistical significance. This finding suggests that RTK/RAS pathway mutations may contribute to disease progression in EOCRC, potentially influencing treatment response. Interestingly, EOCRC NHW patients with RTK/RAS alterations also exhibited a non-significant trend toward reduced survival (p = 0.18), reinforcing the need for larger cohort studies to validate these associations.

The molecular differences identified in this study have significant implications for precision medicine approaches in CRC. The high prevalence of RTK/RAS pathway mutations in EOCRC H/L patients suggests that therapies targeting this pathway, such as MEK inhibitors, may be particularly relevant for this population. However, the presence of NF1 mutations in 11.6% of EOCRC H/L patients raises concerns regarding potential resistance to RAS/MAPK-targeted therapies, highlighting the need for biomarker-driven treatment strategies (13, 14).

The differential Impact of WNT pathway alterations on survival outcomes between NHW and H/L EOCRC patients suggests that ethnicity-specific molecular interactions may influence tumor progression. Future clinical trials should incorporate ethnic diversity in patient recruitment to better assess treatment responses in H/L CRC patients. Additionally, integrating multi-omics approaches—such as transcriptomic and proteomic profiling—will provide deeper insights into the functional consequences of pathway alterations in EOCRC.

Despite its strengths, including the use of large-scale genomic datasets and rigorous statistical analyses, this study has several limitations. First, the retrospective nature of bioinformatics analyses may introduce selection bias, as publicly available genomic databases may not fully represent the broader EOCRC patient population. Second, the relatively small sample size of EOCRC H/L patients may limit statistical power, particularly for survival analyses. Third, this study lacks functional validation experiments, preventing direct mechanistic insights into how specific pathway alterations contribute to tumorigenesis.

To address these limitations, future studies should focus on prospective cohort analyses with larger, more diverse EOCRC patient populations. Additionally, integrating functional genomics approaches will be critical to understanding the biological significance of pathway alterations and their therapeutic implications. Investigating interactions between WNT, TGF-beta, and RTK/RAS pathways will provide a more comprehensive understanding of the molecular drivers of EOCRC, particularly in high-risk populations.

## 5. Conclusion

This study provides novel insights into the molecular heterogeneity of EOCRC in high-risk populations, identifying key ethnic-specific differences in WNT, TGF-beta, and RTK/RAS pathway alterations. H/L patients exhibited higher frequencies of NF1, CBL, MAPK3, and BMPR1A mutations, underscoring the need for further research into their functional roles in tumor progression. While WNT pathway alterations were associated with improved survival in NHW patients, no such benefit was observed in H/L patients, suggesting potential ethnic-specific tumor biology. These findings emphasize the importance of precision oncology approaches that consider ethnic and molecular heterogeneity in EOCRC. By integrating genomic, transcriptomic, and clinical data, future research can refine personalized treatment strategies to improve outcomes and reduce disparities in high-risk populations.

**Figure S1.** Overall survival trends in early-onset Non-Hispanic White (NHW) patients, categorized based on the presence or absence of alterations in the TGF-beta (top left), WNT (top right), and RTK/RAS (bottom) pathways.

**Table S1.** Mutation Frequencies of WNT, TGF-beta, and RTK/RAS Pathway-Associated Genes in Hispanic/Latino Colorectal Cancer Patients Stratified by early-onset colorectal cancer (EOCRC) and late-onset colorectal cancer (LOCRC).

**Table S2.** Alteration frequencies of genes within the WNT, TGF-beta, and RTK/RAS pathways in early-onset colorectal cancer (EOCRC) patients, comparing Hispanic/Latino (H/L) and Non-Hispanic White (NHW) populations.

## Data Availability

All data used in the present study is publicly available at https://www.cbioportal.org/ and https://genie.cbioportal.org. Additional data can be provided upon reasonable request to the authors.

